# Effect of hybrid immunity, school reopening, and the Omicron variant on trajectory of COVID-19 epidemic in India: A modelling study

**DOI:** 10.1101/2022.06.24.22276854

**Authors:** Farhina Mozaffer, Philip Cherian, Sandeep Krishna, Brian Wahl, Gautam I Menon

## Abstract

**Background:** The course of the COVID-19 pandemic has been driven by several dynamic behavioral, immunological, and viral factors. We used mathematical modeling to explore how the concurrent reopening of schools, increasing levels of hybrid immunity, and the emergence of the Omicron variant have affected the trajectory of the pandemic in India, using the model Indian state of Andhra Pradesh (pop: 53 million).

**Methods:** We constructed an age- and contact-structured compartmental model that allows for individuals to proceed through various states depending on whether they have received zero, one, or two doses of the COVID-19 vaccine. Our model is calibrated using results from other models as well as available serosurvey data. The introduction of the Omicron variant is modelled alongside protection gained from hybrid immunity. We predict disease dynamics in the background of hybrid immunity coming from infections and well as an ongoing vaccination program, given prior levels of seropositivity from earlier waves of infection. We describe the consequences of school reopening on cases across different age-bands, as well as the impact of the Omicron (BA.2) variant.

**Results:** We show the existence of an epidemic peak that is strongly related to the value of background seroprevalence. As expected, because children were not vaccinated in India, re-opening schools increases the number of cases in children more than in adults, although most such cases are asymptomatic or mild. The height of this peak reduced as the background infection-induced seropositivity was increased from 20% to 40%. At reported values of seropositivity of 64%, no discernable peak was seen. We also explore counterfactual scenarios regarding the effect of vaccination on hybrid immunity. We find that in the absence of vaccination, even at such high levels of seroprevalence, the emergence of the Omicron variant would have resulted in a large rise in cases across all age bands. We conclude that the presence of high levels of hybrid immunity thus resulted in relatively fewer cases in the Omicron wave than in the Delta wave.

**Interpretation:** In India, the decreasing prevalence of immunologically naïve individuals of all ages helped reduce the number of cases reported once schools were reopened. In addition, hybrid immunity, together with the lower intrinsic severity of disease associated with the Omicron variant, contributed to low reported COVID-19 hospitalizations and deaths.

**Funding:** World Health Organization, Mphasis

## INTRODUCTION

The first confirmed case of COVID-19 was documented on Jan 30, 2020. The initial epidemic wave of COVID-19 in India peaked in September 2020. Thereafter, cases decreased gradually until the middle of February 2021, when a second wave, driven by the emergence of the Delta variant, led to a sharp increase in cases. At the apex of the second epidemic wave, daily reported cases were significantly higher than the previous wave. By late April 2021, India had surpassed 2.5 million active cases, overtaking Brazil as the country with the second highest number of confirmed COVID-19 cases in the world after the US.

Schools have remained mostly closed in India during the pandemic, in an attempt to slow or arrest transmission of the virus.^1^ During the first epidemic wave, school closures together with broader lockdown measures likely contributed to low levels of transmission. However, as other non-pharmaceutical measures were relaxed, the effectiveness of keeping schools closed in India has been widely discussed. Many have argued that the epidemiological benefits of keeping schools closed has been outweighed by the significant impact on cognitive and social development for children. Schools in many states reopened by Aug, 2021, as the Delta wave ended.

India’s COVID-19 vaccination drive began on Jan 16, 2021. Vaccinations were initially targeted at frontline and health-care workers, but the target population was broadened a month later to include the elderly (i.e., >60 years) and those older than 45 years with comorbidities (e.g., diabetes mellitus, coronary heart disease, or hypertension). In later phases, younger age-groups became eligible for vaccines, until all adults older than 18 years were eligible for vaccination by May 1, 2021. From Jan 3, 2022, children between 15 and 18 years have been eligible for COVID-19 vaccination. A third dose was approved for those older than 18 years from April 10, 2022.

COVID-19 vaccines have been found to be extremely efficient in reducing severe cases, hospitalization and death.^2–10^ Mathematical modeling has indicated that vaccinating in an age descending manner^11–13^ is effective at reducing severe illness, hospitalizations, and deaths, even if the vaccine supply is limited. A major challenge with attaining high population vaccination coverage globally, has been vaccine hesitancy in some populations.^14^ Such hesitancy has largely not been significant in India.

The trajectory of COVID-19 around has been marked by the appearance of multiple variants. On Nov 26, 2021, the World Health Organization (WHO) named a new variant of concern B.1.1.529, first discovered in South Africa, as the “Omicron” variant. The first case of Omicron in India was reported on Dec 3, 2021. The BA.2.12.1 Omicron subvariant was the dominant variant in India in early 2022, largely due to its high transmissibility^15–17^ and its immune escape potential.^18–20^ As a result, even settings with high levels of immunity have experienced Omicron-associated epidemic waves.

Attempts to understand the trajectory of the COVID-19 pandemic in India, across multiple waves of the disease, have spurred the development of a number of compartmental models.^12,21–24^ Earlier models were constructed so as to be appropriate to immunologically naïve populations. However, preexisting levels of infection-associated seroprevalence as well as the relatively high population coverage of the vaccination program (e.g., >80), mandate that current models now accommodate additional complexity. Using mathematical models, we aim to answer two specific questions here: (1) why did India not see a large increase in cases following the reopening of schools in many states? and (2) why did the Omicron-associated third epidemic wave not have a significant impact on the population despite higher transmissibility and immune escape potential? We use data from the southern Indian state of Andhra Pradesh to address these two questions here.

## METHODS

### Model

We developed an age-structured compartmental model (Fig 1). Individuals can progress to different compartments based on whether they are unvaccinated, vaccinated with one dose, or vaccinated with two doses. Unvaccinated individuals progress through nine compartments upon infection: Susceptible (S) individuals who are infected move to the Exposed (E) compartment from which they can either become Asymptomatic Infected (IA) and then Recover (R), or become Pre-symptomatic Infected (IP), meaning that they will eventually exhibit symptoms. A certain fraction of Pre-symptomatic individuals become Mildly Infected (IM) before recovering, while the remainder become Severely Infected (IS) and are eventually Hospitalized (H). A fraction of Hospitalized individuals eventually dies, moving to the Decedent (D) compartment, while the remaining recovers.

**Figure 1:**
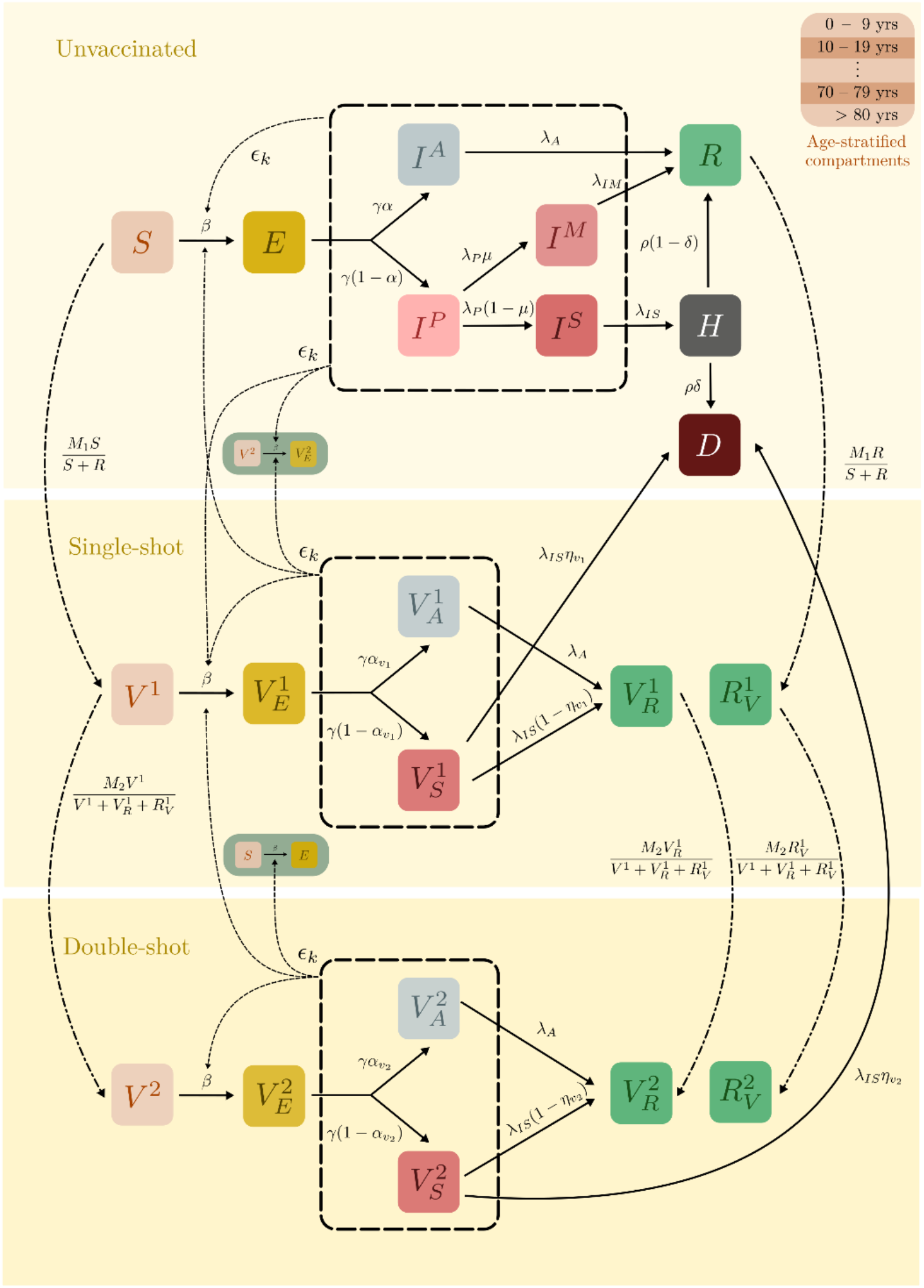
Schematic of model for unvaccinated, one-dose vaccinated, and two-dose vaccinated individuals. Model compartments are each stratified by 10-year age intervals. S = susceptible, E = exposed, IA = asymptomatic infected, IP = pre-symptomatic infected, IM = mildly infected, IS = severely infected, R = recovered, H = hospitalized, D = decedent, V = vaccinated, VE = vaccinated exposed, VA = vaccinated asymptomatic, VS = vaccinated symptomatic, VR = vaccinated recovered, RV = recovered vaccinated. Transition rates and parameters are provided in Tables 1 — 3. To decrease visual clutter, some compartments are shown twice (as small insets). These duplicates correspond to the same single compartment. Dashed lines point to the parameter that is modified by contact matrices and contact parameters. Dash-dotted lines indicate vaccination.

The progression for vaccinated individuals is similar: individuals who have been vaccinated but have not contracted the disease (V) can nevertheless be infected (i.e., breakthrough infection) and move to the Vaccinated Exposed compartment (VE), from which they can either move to the Vaccinated Asymptomatic (VA) or Vaccinated Symptomatic (VS) compartments. The Vaccinated Asymptomatic, as well as a large fraction of the Vaccinated Symptomatic, recover. However, our model also allows for a small fraction of Vaccinated Symptomatic individuals to die depending on age. A distinction is made between individuals who received their vaccine after having contracted the disease (the Recovered Vaccinated; RV) and those who contracted the disease after being vaccinated (the Vaccinated Recovered; VR) to account for the different levels of protection these trajectories might offer, especially where reinfections exist. Separate pathways of compartments exist for both one and two doses of the vaccine. Vaccine efficacy is reflected in the differential rates at which vaccinated individuals can get infected compared to unvaccinated individuals. Additionally, the fraction of infected individuals who progress to symptomatic or severe disease is lower for those in the vaccinated compartments. The parameters governing the disease progression are shown in Tables 1 – 3. We have used parameter values from the INDSCI-SIM model.^25^

**Table 1:**
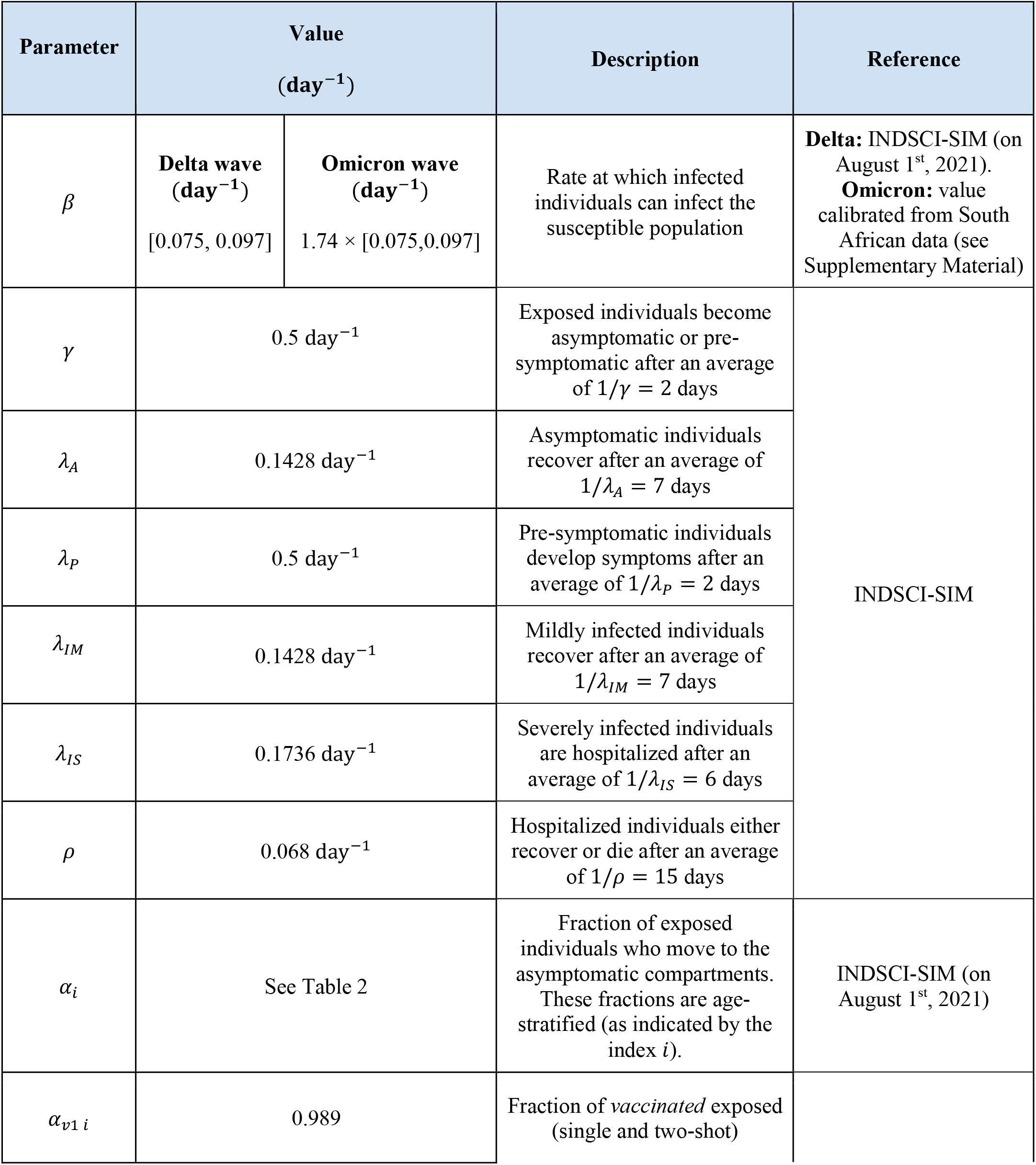
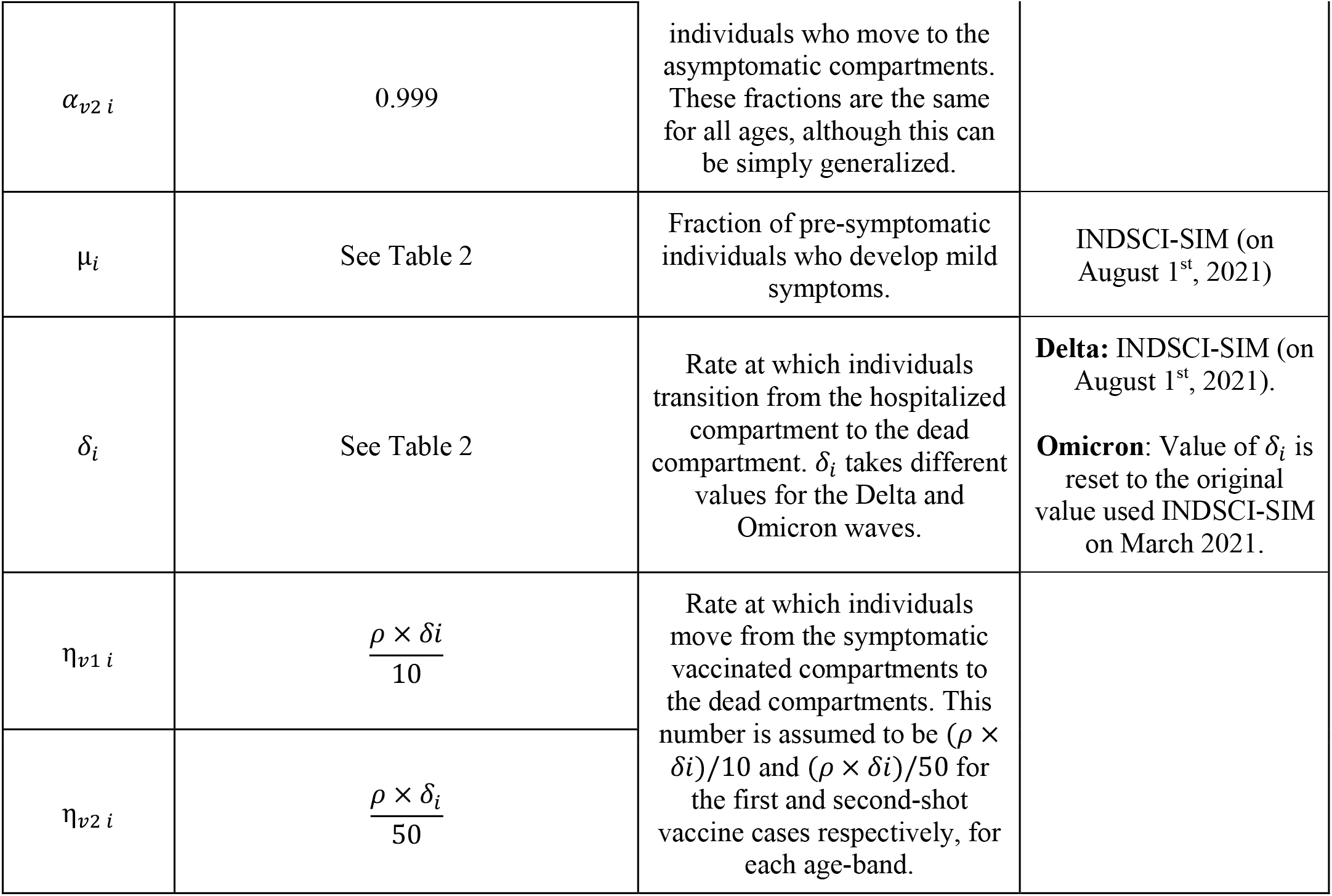
Transition rates and branching parameters for disease progression

| Parameter | Value<br>(day<sup>-1</sup>) |  | Description | Reference |
| --- | --- | --- | --- | --- |
| $\beta$ | **Delta wave**<br>(day<sup>-1</sup>)<br>[0.075, 0.097] | **Omicron wave**<br>(day<sup>-1</sup>)<br>$1.74 \times [0.075, 0.097]$ | Rate at which infected individuals can infect the susceptible population | **Delta:** INDSCI-SIM (on August 1<sup>st</sup>, 2021).<br>**Omicron:** value calibrated from South African data (see Supplementary Material) |
| $\gamma$ | 0.5 day<sup>-1</sup> | | Exposed individuals become asymptomatic or pre-symptomatic after an average of $1/\gamma = 2$ days | INDSCI-SIM |
| $\lambda_A$ | 0.1428 day<sup>-1</sup> | | Asymptomatic individuals recover after an average of $1/\lambda_A = 7$ days | |
| $\lambda_P$ | 0.5 day<sup>-1</sup> | | Pre-symptomatic individuals develop symptoms after an average of $1/\lambda_P = 2$ days | |
| $\lambda_{IM}$ | 0.1428 day<sup>-1</sup> | | Mildly infected individuals recover after an average of $1/\lambda_{IM} = 7$ days | |
| $\lambda_{IS}$ | 0.1736 day<sup>-1</sup> | | Severely infected individuals are hospitalized after an average of $1/\lambda_{IS} = 6$ days | |
| $\rho$ | 0.068 day<sup>-1</sup> | | Hospitalized individuals either recover or die after an average of $1/\rho = 15$ days | |
| $\alpha_i$ | See Table 2 | | Fraction of exposed individuals who move to the asymptomatic compartments. These fractions are age-stratified (as indicated by the index $i$). | INDSCI-SIM (on August 1<sup>st</sup>, 2021) |
| $\alpha_{v1 i}$ | 0.989 | | Fraction of *vaccinated* exposed (single and two-shot) | |

|  |  |  |  |
| --- | --- | --- | --- |
| $\alpha_{v2\ i}$ | 0.999 | individuals who move to the asymptomatic compartments. These fractions are the same for all ages, although this can be simply generalized. | |
| $\mu_i$ | See Table 2 | Fraction of pre-symptomatic individuals who develop mild symptoms. | INDSCI-SIM (on August 1<sup>st</sup>, 2021) |
| $\delta_i$ | See Table 2 | Rate at which individuals transition from the hospitalized compartment to the dead compartment. $\delta_i$ takes different values for the Delta and Omicron waves. | **Delta:** INDSCI-SIM (on August 1<sup>st</sup>, 2021).<br>**Omicron:** Value of $\delta_i$ is reset to the original value used INDSCI-SIM on March 2021. |
| $\eta_{v1\ i}$ | $\frac{\rho \times \delta_i}{10}$ | Rate at which individuals move from the symptomatic vaccinated compartments to the dead compartments. This number is assumed to be $(\rho \times \delta_i)/10$ and $(\rho \times \delta_i)/50$ for the first and second-shot vaccine cases respectively, for each age-band. | |
| $\eta_{v2\ i}$ | $\frac{\rho \times \delta_i}{50}$ | | |

**Table 2:**
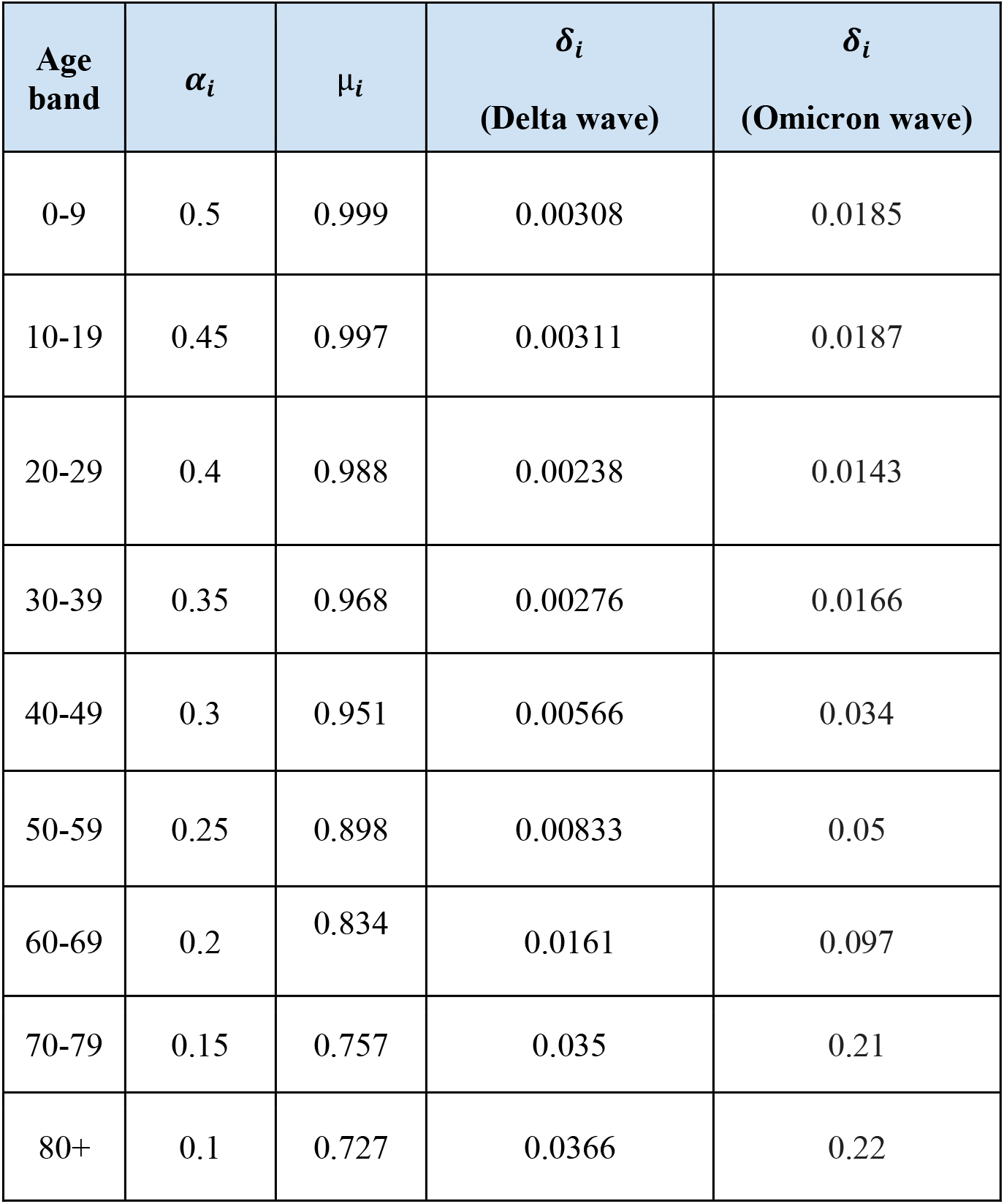
Age-stratified branching parameters for disease progression

**Table 3:**
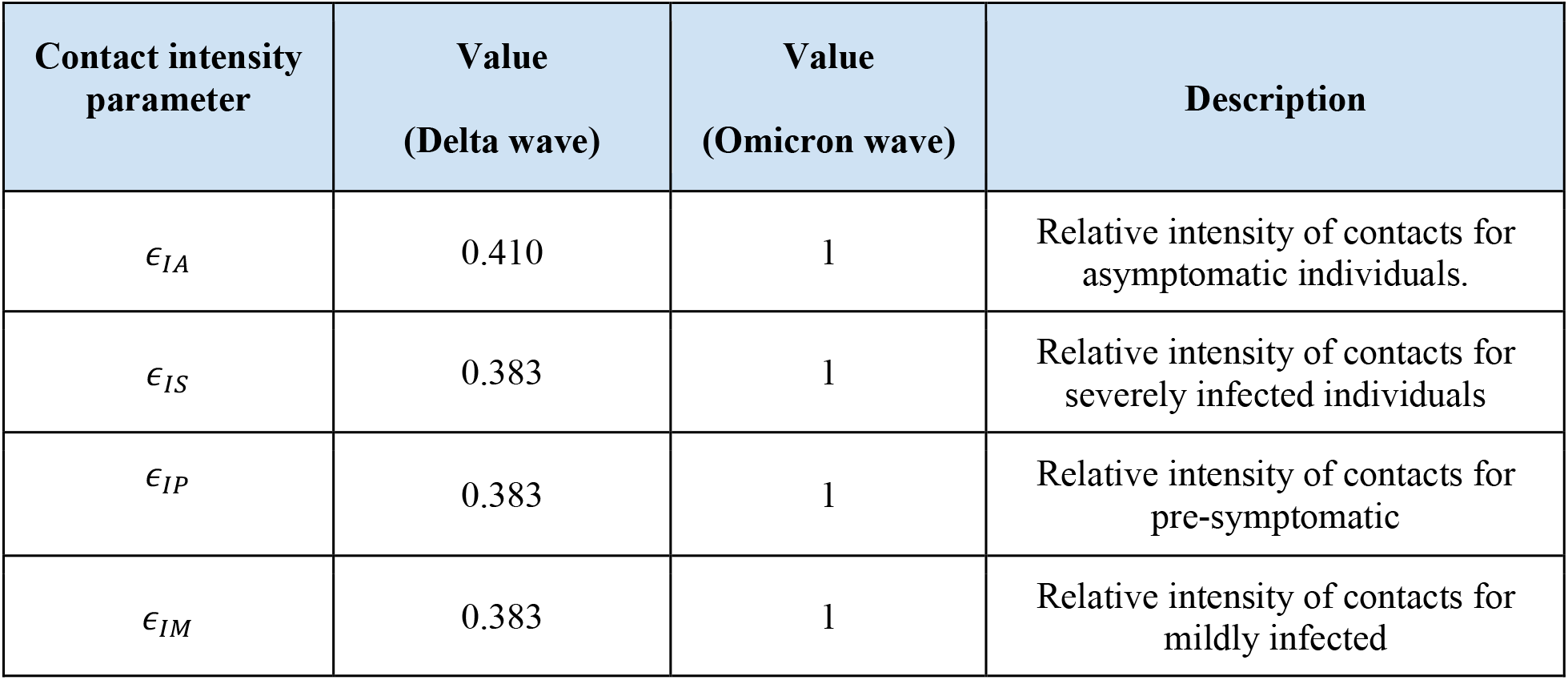
Contact parameters that modulate the interactions between susceptible and infected individuals

| Contact intensity parameter | Value (Delta wave) | Value (Omicron wave) | Description |
| --- | --- | --- | --- |
| $\epsilon_{IA}$ | 0.410 | 1 | Relative intensity of contacts for asymptomatic individuals. |
| $\epsilon_{IS}$ | 0.383 | 1 | Relative intensity of contacts for severely infected individuals |
| $\epsilon_{IP}$ | 0.383 | 1 | Relative intensity of contacts for pre-symptomatic |
| $\epsilon_{IM}$ | 0.383 | 1 | Relative intensity of contacts for mildly infected |

The population is divided into age bands grouped into 10-year intervals: 0-9, 10-19, 20-29, 30-39, 40-49, 50-59, 60-69, 70-79, and 80+ years. Contacts between different age bands are differential, reflecting the social structure of the population. An age-specific contact matrix is used to account for the impact of non-pharmaceutical interventions, including lockdowns and school closures. The contact matrix *C*_*ij*_ ultimately governs the force of infection on each age-band *i* due to the age-band *j*.

The parameter *β*, which governs the force of infection for every susceptible individual, is assumed to be the same for all individuals, independent of their vaccination status. The value of *β* was obtained from simulations of the INDSCI-SIM model, where it is estimated using a Bayesian approach to match daily reported cases and deaths of Andhra Pradesh. This approach gives us a likely value of *β*, as well as a confidence interval. Our simulations are run for a range of *β* within this interval, providing us with a band of potential disease trajectories. In addition, this quantity is further modulated by the relative intensity of contacts between susceptible and different kinds of infectious individuals whose effect is simply specified here through factors of *ϵ*. These values are obtained from the INDSCI-SIM simulations, and are held constant throughout the Delta wave. At the onset of Omicron, these values are reset to 1, indicating that both asymptomatic and symptomatic individuals are equally likely to infect susceptible individuals during the Omicron wave.

To estimate the undercounting of daily cases, a bias factor of 11 and 22 is used for Andhra Pradesh before and after the Omicron wave, respectively. The first factor is taken from the INDSCI-SIM predictions for July 2021, while the second is an assumption based on the reduced levels of testing in the Omicron wave, where a good fraction of the population chose to remain untested or their test results were not reported, as home-based rapid antigen tests were more accessible to much of the population.^26–28^

### Initial conditions

We begin all simulations of our model from August 1, 2021. The initial conditions for our simulation were obtained by running the INDSCI-SIM model from March 1, 2020, to Aug 1, 2021. Values in each compartment were then used as the initial condition for our vaccination model. India’s vaccination drive began on Jan 16, 2021. From March 1, 2020 individuals above older than 60 years and individuals older than 45 years with comorbidities were eligible for vaccination, and on the 1st of April, this was relaxed to include all individuals above the age of 45. On the 1st of May, all individuals above the age of 18 were eligible for vaccination. Since the original INDSCI-SIM model did not implement vaccinations, it was necessary to estimate the number of individuals who had been vaccinated with either one or two doses. These numbers are obtained from covid19india.org.

The INDSCI-SIM simulation gave us a background infection-induced seropositivity of roughly 64%, close to the actual seropositivity measured in the serosurvey conducted from June to July, 2021.^29^ When we use the term “seropositivity” in this paper to refer to our initial condition on Aug 1, 2021, we refer to the population prevalence of antibodies induced by infection, rather than both from infection and vaccination, a reasonable approximation at low levels of vaccination.

In order to study the effect of different initial seropositivity levels, we also conducted simulations in which we adjusted the number of individuals in the susceptible and recovered compartments so that the total number of recovered matched the infection-induced seropositivity levels for which we aimed. For example, for seropositivity levels lower than 64%, an appropriate number of individuals was transferred from the recovered to the susceptible compartment, while the inverse was done for seropositivity levels greater than 64%.

A constant fraction of the susceptible and recovered individuals, based on the vaccine coverage in our study region, were transferred from the Susceptible compartment to the Vaccinated (V1, V2) and Recovered Vaccinated (RV1, RV2) compartments, representing the fraction of people who had received vaccines by the simulation start date. These numbers were then divided among the first (V1, RV1) and second dose (V2, RV2) vaccine compartments based on the details of the vaccine coverage by the date of the simulation’s start. For example, 30% of the population of the state of Andhra Pradesh was vaccinated with at least one dose, and 10% with the second dose, by the Aug 1, 2021. We thus move 20% of all susceptible individuals to the V1 compartment and 10% to the V2 compartment. This is repeated for the individuals in the recovered compartment to the RV1 and RV2 compartments respectively. We ignore the number of individuals who were infected at the time of vaccination, as we expect this number to be very low.

### Reopening schools

Our simulation, like the INDSCI-SIM model, uses contact matrices to model the contacts between individuals of different age-bands.^30^ In our simulations, we consider all individuals below the age of 20 to be children. This age-band (technically 0-18, but we ignore this minor discrepancy) is not eligible to receive vaccines. The remaining population is considered to be adult. The contact matrix method allows us to model the effects of social-structures such as those arising from household and work contacts. This method can therefore also be used to model the effects of non-pharmaceutical interventions like lockdowns and restrictions on public transport. In particular, these contact matrices (shown in Table 4) can be used to model the reopening of schools, by introducing more contacts between the children, as well as between children and adults.

**Table 4:** Weights used for age-specific contact matrices Weights used to create the contact matrices that simulate social contacts and mixing within the population. Four different locations (Home, School, Work, and Others) were used to address the contacts between different age groups. At home all age groups have contacts with each other with equal weights. The same is true for the “Others” location, but with half the weight as the Home location. In schools, children in the below 20 age-bands have high contact with each other, while teachers (age-bands 20 and above) have moderate contact with each other as well as with the children. See Prem et al. and Hazra et al. for more details.

| Model | Home | School | Work | Others |
| --- | --- | --- | --- | --- |
| INDSCI-SIM model<br>(from March, 2020 to 31-07-2021) | 1 | 0 | 0.5 | 0 |
| Hybrid immunity model<br>(Before school open, 01-08-2021 to 15-08-2021) | 1 | 0 | 0.5 | 0.5 |
| Hybrid immunity model<br>(After school open, 16-08-2021 to 15-12-2021) | 1 | 0 to 0.5 | 0.5 | 0.5 |
| Hybrid immunity model<br>(Omicron, South Africa) | 1 | 0.5 | 1 | 0.5 |
| Hybrid immunity model<br>(Omicron, Andhra Pradesh & district, from 16-12-2021) | 1 | 0 | 0.5 | 0.5 |

We use contact matrices that are a weighted average over the contributions from different types of “locations” (e.g., home, workplace, school).^25,30^ The weights may vary with time, appropriate to the interventions implemented at that point in time, and are described in Table 4.

In the INDSCI-SIM simulation, which we use to obtain the initial conditions for the hybrid-immunity model, we assume that schools are closed, and that social interactions due to public transport and other social gatherings are minimized. As a result, the contribution to the contact matrices of household contacts is 1 and work contacts is 0.5. Both school and public gathering contacts were assumed to be 0. In the hybrid immunity model proposed in this paper, we allow for the existence of many more interactions. We assume the existence of “other” social interactions like public transport and social gatherings, and add a contribution of 0.5 to the contact matrices for the first fifteen days, while keeping schools closed. Home and work contact contributions were left unchanged. These choices can of course be varied to represent other intervention scenarios

Reopening schools was modeled by gradually increasing the corresponding values of the contact matrices in intervals of 20 days. School interactions were thus ramped up from 0% to 50% over roughly 120 days, which corresponds to the Dec 15, 2021. This coincides with the onset of the Omicron wave in India, and therefore, we further assume all schools to be closed from Dec 15, 2021 until the end of the simulation in March, 2022.

### Emergence of Omicron variant

The Omicron variant of concern was first detected in South Africa, before spreading to more than 90 countries. To describe the epidemiology of the Omicron variant, we calibrated our model with data from South Africa (See Supplementary Material for more information), using daily reported cases to estimate its growth rate.

We simulated the growth of cases associated with the Omicron variant in South Africa from Nov 15, 2021. Given that by then, the total vaccinated population in South Africa was 27% and 22% for the first and second doses, respectively, we use this input in our calculations. We initially assume a seropositivity of 60% and assume that the effects of a prior infection can be taken to be equivalent to that of a single vaccination, 20% of the population from the recovered compartment were moved to the V1 compartment from where they can be infected.

The weight factors multiplying the home and work contact matrices were taken to be 1, while the weight factors of schools and “other” were taken to be 0.5, assuming that no strict restrictions were implemented in South Africa during the Omicron wave. The population was vaccinated at a low vaccination rate. All the parameters for the unvaccinated except *β* are the same as used in the INDSCI-SIM model. We have chosen a value of *β* such that the increase in number of daily cases matches the reported daily cases. To account for case-undercounting, a “bias” factor of 15 is used to best fit the South African data for cases and deaths accounting for the fact that many symptomatic patients chose not to be tested. This factor was further optimized to fit the Indian data, based on the expected seroprevalence and the test positivity.

### Vaccine allocation

The rate at which individuals receive the first vaccine dose depends only on the number of individuals eligible for a first dose. The same is true for the second dose. Additionally, any excess first doses are used to vaccinate the individuals eligible for second doses. The same process is repeated for the second dose, with any excess doses being used to vaccinate eligible unvaccinated people. This process ensured that all available vaccines were used, whenever possible. The details of our calculations can be found in the Supplementary Material.

We study the possible outcomes for different background seroprevalence levels of 20%, 40%, and 80% in order to assess the importance of hybrid immunity.

## RESULTS

Figure 2 shows the daily number of cases, aggregated over all age-bands for Andhra Pradesh, along with the three other scenarios involving different background seroprevalence levels. The results for adults and children can be found in the Supplementary Material.

**Figure 2:**
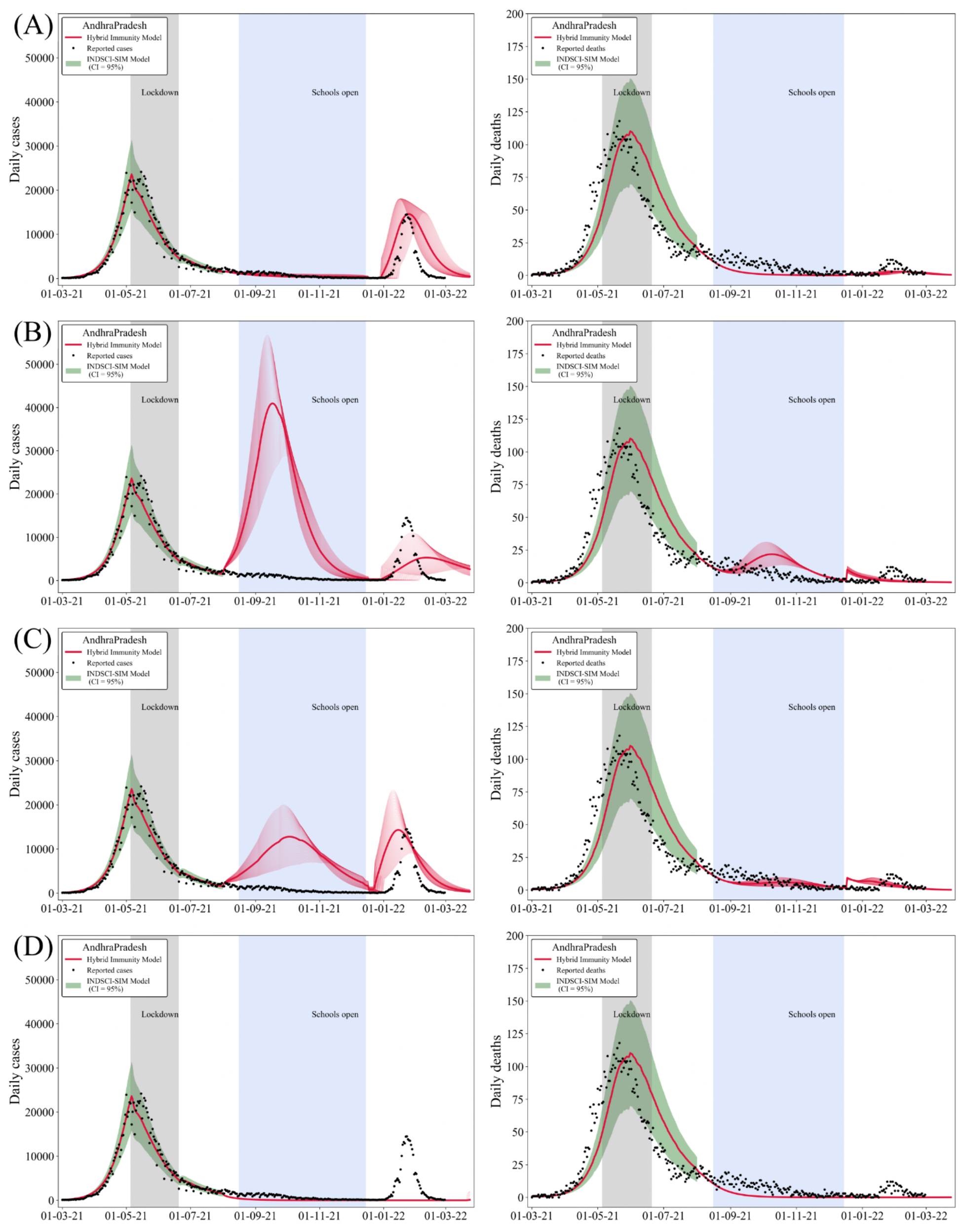
Effect of infection-induced seropositivity on school reopening. The daily number of cases (left panel) and the daily number of recorded deaths (right panel) for all age-groups, for the entire state of Andhra Pradesh, as well as three scenarios that demonstrate a variation in background seroprevalence. The top-most panel represents a background seroprevalence of 64%, while the remaining are for 20%, 40%, and 80% seroprevalence respectively. The black dots represent the recorded daily numbers obtained from covid19bharat.org, while the confidence intervals are obtained from the INDSCI-SIM simulation. The initial part of the graph follows the INDSCI-SIM prediction until the 1st of August. Schools are gradually opened, as described in the Methods section, over a period of 120 days. The numbers are scaled by an appropriate bias factor to account for daily undercounting of cases.

Figure 2A shows that the absence of a noticeable rise in the daily number of cases or deaths, in both older and younger age-bands when schools were reopened, can be explained by the fact that the background seroprevalence was sufficiently high.

For example, had the seroprevalence been lower (e.g., 20%) we could have expected to see a sharp rise in cases with a high peak around the end of September, as can be seen in the Figure 2B. Our simulations indicate that this peak would have affected children, who were unvaccinated, much more than adults. For example, in the case where the seropositivity was 20%, it was found that the height of the school reopening peak relative to the Delta wave’s peak was roughly 2.8 times for children, while it was only 1.3 times for adults. If the background seroprevalence had been close to 40%, the height of the school reopening peak would have been around 0.9 times the Delta peak for children, while the height relative to the Delta peak would have only been 0.33 times for adults.

Once the seroprevalence crosses 60%, the school-reopening-associated epidemic peak is completely washed out, indicating that the level of immunity attained through the combination of both vaccination and prior infection is sufficient to curtail the spread of the disease. Even when the seroprevalence was 20%, while a sharp rise in daily cases was observed, these cases were mostly asymptomatic or mild, with the number of severe cases remaining quite low in comparison to numbers in the Delta wave. Our simulations found these cases to occur mainly among the adult population who had more of a chance of severe illness than children.

The introduction of Omicron – modeled by an increase in *β* which was calibrated by the data from South Africa – causes an upswing in the number of cases across all age-bands, as expected. As described in the Methods section, the possibility of reinfection by Omicron is modeled by moving a constant fraction of 20% of the recovered individuals to the “single-dose-vaccinated’’ compartment V1. The locations of the peak of the Omicron wave in our simulations match actual data reasonably well. However, the duration of the wave in our simulations appears to be larger. A larger value of *β* would have allowed us to match the observed width.

Figure 3 shows the counterfactual scenario where our simulation was run for a seropositivity of 64% (as in Figure 2A) but without incorporating the effects of vaccination. As can be seen, while this would not have affected school reopening, a much higher peak could have been expected during the Omicron wave. Both children and adults would have fared similarly during this wave, as is to be expected given that both groups are now unvaccinated. This would have resulted in a rise in the number of severe cases (and, consequently, deaths) over all age-bands that would have been much larger than the numbers observed during the Delta wave.

**Figure 3:**
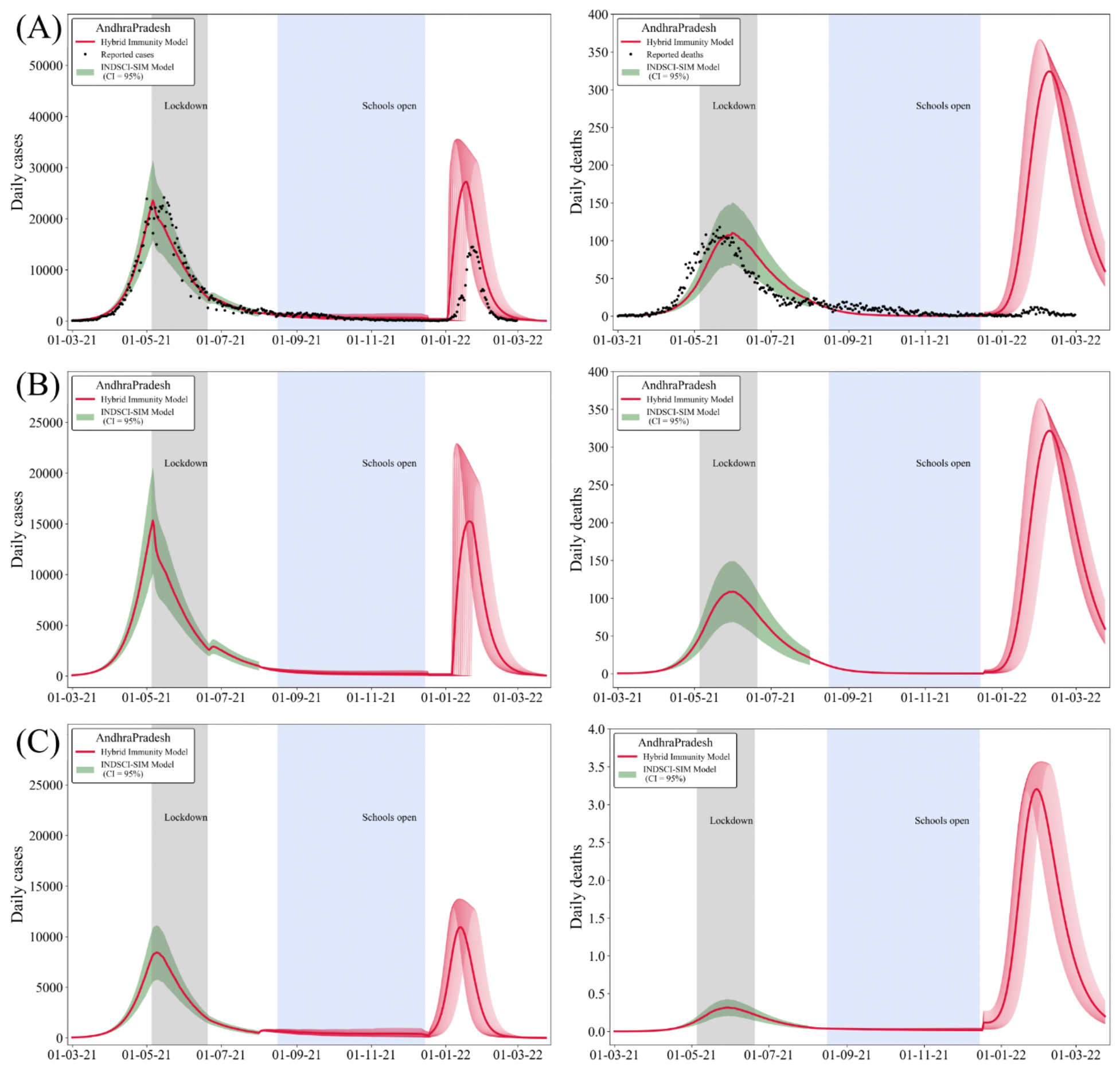
Role of vaccination in hybrid immunity. Results of a simulation with no vaccination, to quantify the effect that vaccination has on hybrid immunity. All panels represent results for a seropositivity of 64%, close to the reported value from serosurvey data. The panels on the left show the daily number of cases, while those on the right show the daily number of deaths. (A) shows the results for all age groups, (B) the results for adults (above 20) and (C) the results for children (below 20). Our simulations indicate that a much higher Omicron peak could have been expected if a smaller fraction of the population had been vaccinated, as can be seen by comparing Figures 3A and 2A.

## DISCUSSION

Ours may be the first model to incorporate hybrid immunity into discussions of the COVID-19 trajectory of India, and to assess its specific impact on school reopening and the Omicron case trajectory. The model contains multiple interacting time-varying components, all of which are essential to understanding the dynamics of the disease across the Delta and Omicron waves. Our model includes a two-dose vaccine and differential levels of protection from these doses, across different age-bands.

Our results explain why cases from the Delta wave continued to decline even as schools were reopened across the state of Andhra Pradesh. Given that seropositivities in adults and children are comparable (from ICMR surveys at the end of the second wave), we conclude that the impact of school reopening on case-load was substantially reduced by the fact that a large fraction of the population had already been infected. Our simulations indicate that at the levels of seropositivity at which schools were reopened, vaccine coverage played a subdominant role in curtailing the spread of the disease, when compared to the role played by the background infection-induced seropositivity. We show, through model calculations at different levels of seropositivity, that low seropositivity values would have led to a much more substantial effect on infections in school-going children.

Next, we showed that the impact of the Omicron trajectory was reduced in India for two reasons: the high background seropositivity from infection and the high vaccine coverage, in addition to the overall reduced severity of disease caused by this variant. While we chose conservative values for the extent of reinfections as well as for break-through infections, our model can easily account for larger numbers if other, more aggressively immune-evading variants, were to emerge.

The lessons we draw from our work are the following. At large background seropositivity levels, especially if background prior infection levels in children are comparable to those in adults, schools can and should be reopened. This should have only a marginal impact on cases. Further, our model suggests that increased immune evasion leading to an effectively larger susceptible population can lead to cases increasing sharply, indicating that the immune escape potential of new variants should be carefully tracked. Finally, given that variants of the original Omicron strain appear to be dominant across the world currently, the development of new vaccines that target Omicron variants should be a priority.

We conclude that Andhra Pradesh did not see a large rise in cases among the unvaccinated after schools began reopening because of high background levels of seropositivity. To further address this claim, we experimented with various counterfactual scenarios to show that a rise in cases would have been expected with lower levels of prior seropositivity. Our discussion of the Omicron wave illustrates that even with its higher transmissibility, the impact of this variant was blunted by high levels of hybrid immunity. We verify this through the construction of a number of counterfactual scenarios where we can alter the balance of infection- and vaccination-induced seroprevalence.

## CONTRIBUTORS

FM, PC, and BW did the literature search. BW, SK, and GIM designed the study. Analyses were interpreted by all authors. FM and PC wrote the original draft of the manuscript which was reviewed and edited by all other authors (SK, BW, and GIM). All authors (FM, PC, SK, BW, and GIM) had full access to all the data. BW and GIM had final responsibility for the decision to submit for publication.

## Data Availability

Aggregated case and vaccination data are available www.covid19india.org and www.covid19bharat.org.

https://www.covid19india.org

## DATA SHARING

Aggregated case and vaccination data are available www.covid19india.org and www.covid19bharat.org.

## ACKNOWLEDGEMENTS

The authors acknowledge support from the World Health Organization SAGE Working Group on Vaccines (APW Contract #202706833), Mphasis, and the Centre for Bioinformatics and Computational Biology at Ashoka University. SK acknowledges funding from the Department of Atomic Energy, Government of India (RTI 4006), the Simons Foundation (287975) and the Science & Engineering Research Board, Department of Science & Technology, Government of India (MTR/2020/000253). We acknowledge discussions with the Andhra Pradesh Epidemiological Committee and in particular with Profs. Gagandeep Kang and Jacob John. We are grateful to Dhiraj Kumar Hazra for many discussions concerning the use of INDSCI-SIM.

