## supplemental material for "Effect of hybrid immunity, school reopening, and the Omicron variant on trajectory of COVID-19 epidemic in India: A modelling study"

### APPENDIX

#### A1. Model Formulation

The dynamics of the progression through the compartments is governed by the equations given below. We define the total number of infectious individuals in age band *j* that might infect susceptible individuals (i.e., the total number of individuals from age band *j* in all infected compartments except hospitalized.) by the quantity $I_j^{\text{eff}}$.

$$I_j^{\text{eff}} = \frac{\epsilon_{IP} I_j^P + \epsilon_{IA} I_j^A + \epsilon_{IM} I_j^M + \epsilon_{IS} I_j^S + \epsilon_{IA} V_j^{1A} + \epsilon_{IS} V_j^{1S} + \epsilon_{IA} V_j^{2A} + \epsilon_{IS} V_j^{2S}}{N_j}$$

The rates of the different transitions are given in Tables 1—3. $M_1$ and $M_2$ in the below equations are the total rates at which the first and second vaccine doses are administered across all eligible compartments. The mathematical form of the vaccination rates is explained in the section "Vaccine dose allotment".

$$\frac{dS_i}{dt} = -\ \beta S_i \sum_{j=1}^{N_{\text{age}}} C_{ij}\, I_j^{\text{eff}} - M_1 \left(\frac{S_i}{S_i + R_i}\right)$$

$$\frac{dE_i}{dt} = \beta S_i \sum_{j=1}^{N_{\text{age}}} C_{ij}\, I_j^{\text{eff}} - \gamma E_i$$

$$\frac{dI_i^A}{dt} = \alpha\gamma E_i - \lambda_A I_i^A$$

$$\frac{dI_i^P}{dt} = (1-\alpha)\gamma E_i - \lambda_P I_i^P$$

$$\frac{dI_i^M}{dt} = \mu\lambda_P I_i^P - \lambda_{IM} I_i^M$$

$$\frac{dI_i^S}{dt} = (1-\mu)\lambda_P I_i^P - \lambda_{IS} I_i^S$$

$$\frac{dH_i}{dt} = \lambda_{IS} I_i^S - \rho H_i$$

$$\frac{dR_i}{dt} = (1-\delta)\rho H_i + \lambda_A I_i^A + \lambda_{IM} I_i^M - M_1 \left(\frac{R_i}{S_i + R_i}\right)$$

$$\frac{dD_i}{dt} = \delta\rho H_i + \lambda_{IS}\eta_{v1} V_i^{1S} + \lambda_{IS}\eta_{v2} V_i^{2S}$$

$$\frac{dV_i^1}{dt} = -\beta V_i^1 \sum_{j=1}^{N_{\text{age}}} C_{ij} I_j^{\text{eff}} - M_2\left(\frac{V_i^1}{V_i^1+V_i^{1R}+R_i^{1V}}\right) + M_1\left(\frac{S_i}{S_i+R_i}\right)$$

$$\frac{dV_i^{1E}}{dt} = \beta V_i^1 \sum_{j=1}^{N_{\text{age}}} C_{ij} I_j^{\text{eff}} - \gamma V_i^{1E}$$

$$\frac{dV_i^{1A}}{dt} = \alpha_{v1}\gamma V_i^{1E} - \lambda_A V_i^{1A}$$

$$\frac{dV_i^{1S}}{dt} = (1-\alpha_{v1})\gamma V_i^{1E} - \lambda_{IS} V_i^{1S}$$

$$\frac{dV_i^{1R}}{dt} = \lambda_{IS}(1-\eta_{v1}) V_i^{1S} + \lambda_A V_i^{1A} - M_2\left(\frac{V_i^{1R}}{V_i^1+V_i^{1R}+R_i^{1V}}\right)$$

$$\frac{dR_i^{1V}}{dt} = M_1\left(\frac{R_i}{S_i+R_i}\right) - M_2\left(\frac{R_i^{1V}}{V_i^1+V_i^{1R}+R_i^{1V}}\right)$$

$$\frac{dV_i^2}{dt} = -\beta V_i^2 \sum_{j=1}^{N_{\text{age}}} C_{ij} I_j^{\text{eff}} + M_2\left(\frac{V_i^1}{V_i^1+V_i^{1R}+R_i^{1V}}\right)$$

$$\frac{dV_i^{2E}}{dt} = \beta V_i^2 \sum_{j=1}^{N_{\text{age}}} C_{ij} I_j^{\text{eff}} - \gamma V_i^{2E}$$

$$\frac{dV_i^{2A}}{dt} = \alpha_{v2}\gamma V_i^{2E} - \lambda_A V_i^{2A}$$

$$\frac{dV_i^{2S}}{dt} = (1-\alpha_{v2})\gamma V_i^{2E} - \lambda_{IS} V_i^{2S}$$

$$\frac{dV_i^{2R}}{dt} = \lambda_{IS}(1-\eta_{v2}) V_i^{2S} + \lambda_A V_i^{2A} + M_2\left(\frac{V_i^{1R}}{V_i^1+V_i^{1R}+R_i^{1V}}\right)$$

$$\frac{dR_i^{2V}}{dt} = M_2\left(\frac{R_i^{1V}}{V_i^1+V_i^{1R}+R_i^{1V}}\right)$$

### A2. Vaccine Dose Distribution

Vaccine doses were allocated while keeping two things in mind: (i) The doses should be assigned per age-band according to the number of eligible individuals in that age-band, and (ii) The doses should be distributed with some fraction being first (and some fraction second) doses. However, dose wastage should be minimized.

#### Distribution of first doses

Let us assume that there is some number $D_F$ of first doses, and we need to distribute this number amongst the population. For simplicity, let us consider the case where there are only two age-bands. This analysis can easily be generalized to a larger number of bands. Let the eligible population in each band be denoted by $E_1$ and $E_2$ respectively. We represent the age-stratified population as a vector, with each component representing the number of individuals in each age-band. In this case, the doses $D_F$ will be distributed amongst the two age-bands as:

$$\text{First doses per age-group per day} = D_F \begin{pmatrix} \frac{E_1}{E_1+E_2} \\ \frac{E_2}{E_1+E_2} \end{pmatrix},$$

where each component of the above vector represents the number of doses to be distributed to the first and second age-bands respectively. In this case, the number of people eligible for vaccines are the number of susceptible and the number of recovered individuals in the population at that instant of time. In other words,

$$E_1 = S_1 + R_1$$

$$E_2 = S_2 + R_2$$

Therefore,

$$\text{First doses per age-group per day} = \frac{D_F}{(S_1+R_1) + (S_2+R_2)} \begin{pmatrix} S_1 + R_1 \\ S_2 + R_2 \end{pmatrix}$$

#### Distribution of second doses

Let's assume a number $D_S$ of second doses. The same argument as before holds, the only difference is what constitutes the number of "eligible" people. In this case, the number of eligible people for the second dose are those in the Vaccinated, Vaccinated Recovered, and Recovered Vaccination compartments (for a single shot). In other words,

$$E_1 = V_1^{(1)} + V_{R1}^{(1)} + R_{V1}^{(1)},$$
$$E_2 = V_2^{(1)} + V_{R2}^{(1)} + R_{V2}^{(1)},$$

where, for example, the term $V_{R2}^{(1)}$ represents the number of one-shot vaccinated recovered individuals after the first shot in age-group 2. Thus, similar to before,

$$\text{Second doses per age-group per day} = \frac{D_S}{(V_1^{(1)}+V_{R1}^{(1)}+R_{V1}^{(1)})+(V_2^{(1)}+V_{R2}^{(1)}+R_{V2}^{(1)})} \begin{pmatrix} V_1^{(1)} + V_{R1}^{(1)} + R_{V1}^{(1)} \\ V_2^{(1)} + V_{R2}^{(1)} + R_{V2}^{(1)} \end{pmatrix}$$

The above distribution of doses makes sure that we distribute the vaccines proportionally across different age-bands, based on the number of eligible individuals.

**Minimizing vaccine wastage**

The numbers $D_F$ and $D_S$ are not only set by the daily vaccination rate, but are also chosen so as to minimize vaccine wastage. The idea is to split the assignment of doses into two stages: in the first stage, everyone who is eligible gets their first or second shot according to the some assigned numbers $D_1$ and $D_2$, given by the daily vaccination rate for first and second doses ($M_1$ and $M_2$) respectively. In principle, there could be cases where certain individuals eligible for a first dose do not get one, despite there being a surplus of second doses (or vice versa). To address this, in the second stage, any extra doses that exist are distributed among the individual remaining eligible (but unvaccinated) individuals.

The algorithm we used is as follows: We start of by finding the number of unused first doses. This is the difference between the total number of assigned "first" doses and the total number of eligible people. We also compute the number of unused second doses in the same fashion. Then, we can find the number of unused first and second doses using:

$$\text{Unused first doses} \quad = U_1 = \max\left(D_1 - \left(E_1^{(1)} + E_2^{(1)}\right) \;,\; 0\right)$$
$$\text{Unused second doses} = U_2 = \max\left(D_2 - \left(E_1^{(2)} + E_2^{(2)}\right) \;,\; 0\right)$$

Next, we give all the unused first doses to those who are eligible for a second dose but haven't received one, and vice versa. Of course, the total number of doses administered can never be greater than the total number of eligible people. As a result, we have

First doses finally administered $\quad D_F = \min\left(\min\left(D_1 \;,\; E_1^{(1)} + E_2^{(1)}\right) + U_2 \;,\; E_1^{(1)} + E_2^{(1)}\right)$

Second doses finally administered $\quad D_S = \min\left(\min\left(D_2 \;,\; E_1^{(2)} + E_2^{(2)}\right) + U_1 \;,\; E_1^{(2)} + E_2^{(2)}\right)$

**A3. Calibration for Omicron variant**

Figure A3.1: **Calibration from South African data**

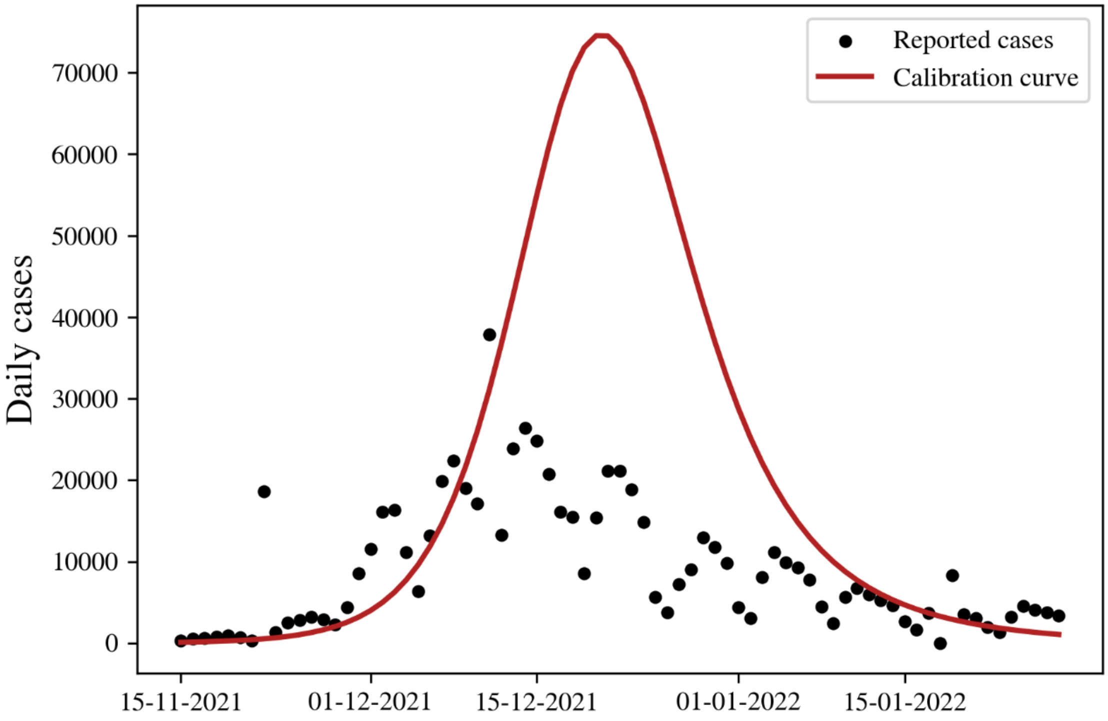


Daily cases during the Omicron wave in South Africa: Black circles represent the daily reported cases,[1] while the red curve is a calibration curve used to match the initial rise in reported cases.

We used our model on the data shown in Figure (A3.1) to match the initial spread of the disease by varying the value of $\beta$. A 1.74-fold increase in $\beta$ from the initial value used to fit cases for the Delta variant in Andhra Pradesh was found to match the initial spread. Additionally, a constant bias factor of 15 was used to model the undercounting of cases.

Later in the Omicron wave, those infected may have chosen to simply not get tested given the overall mildness of the disease caused by this variant.[2,3] We believe this accounts for the discrepancy between our calibration curve and the reported data.

### A4. Age-stratified results for adults and children

Figure A4.1: **Effect of infection-induced seropositivity and school reopening on adults**

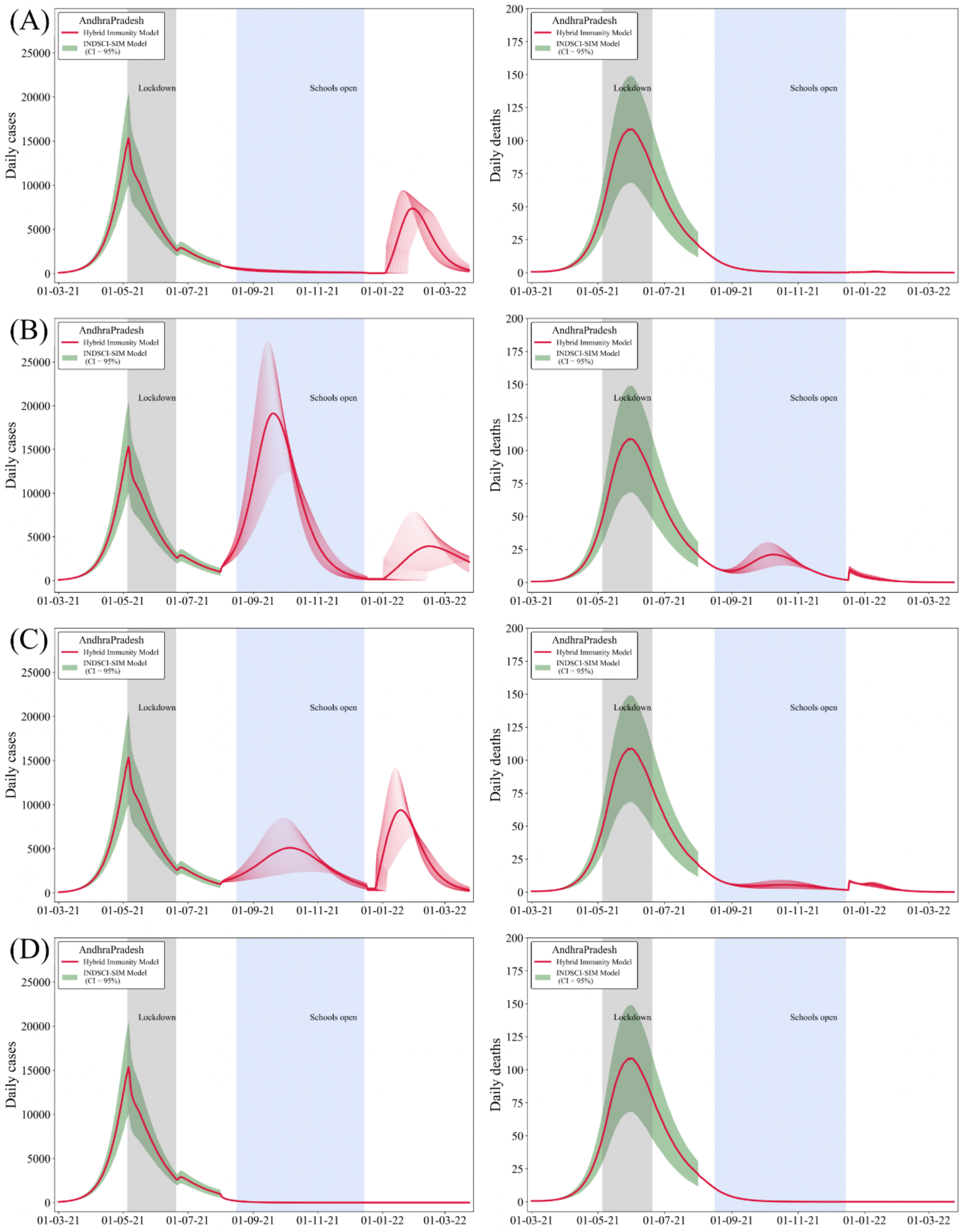


The daily number of cases (left panels) and the daily number of recorded deaths (right panels) for the same study areas as in Figure (2) in the main text, but restricted only to adults (i.e. above the age of 20).

Figure A4.2: **Effect of infection-induced seropositivity and school reopening on children**

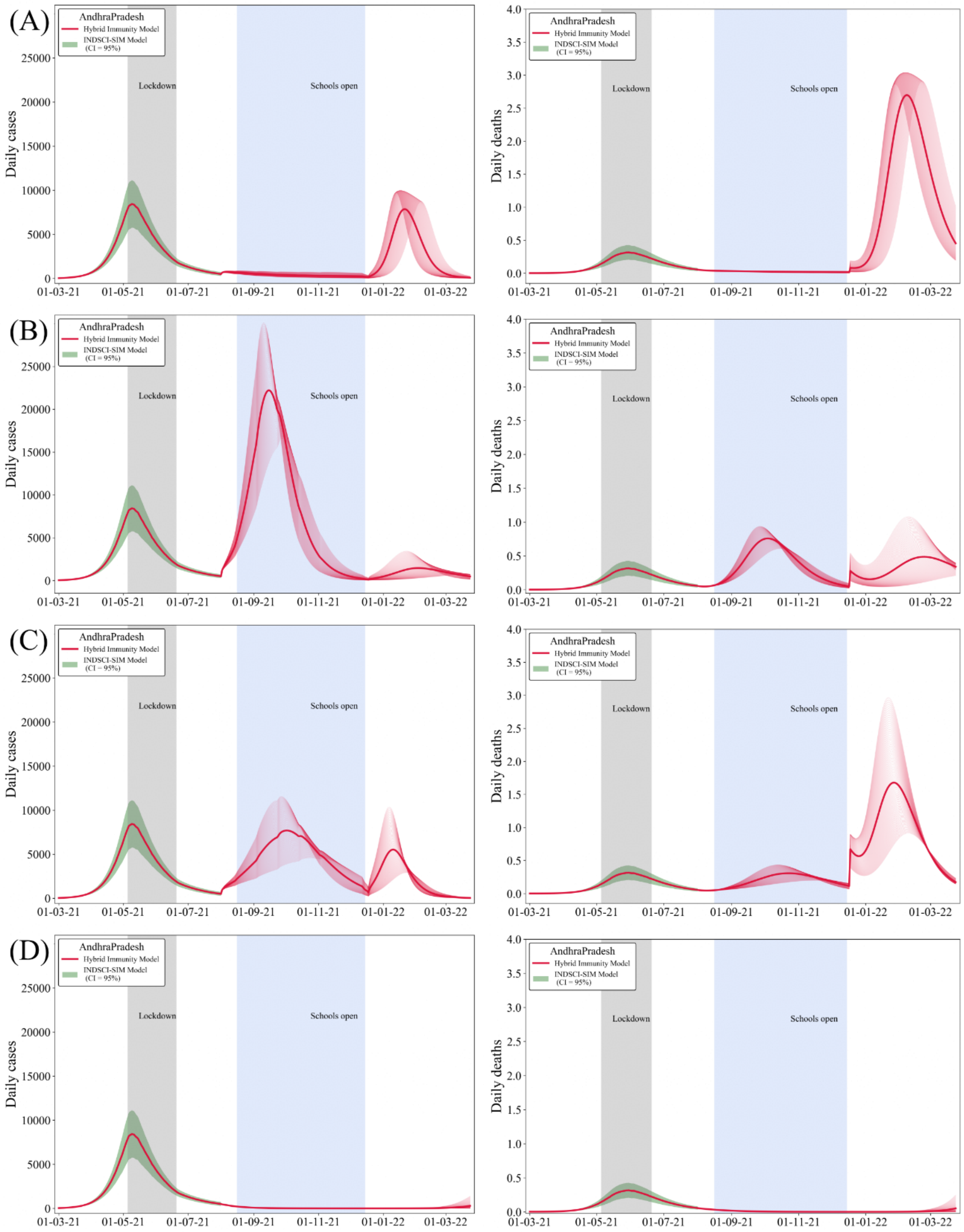


The daily number of cases (left panels) and the daily number of recorded deaths (right panels) for the same study areas as in Figure (2) in the main text, but restricted only to children (i.e. individuals below the age of 20).
